# Determinants of Job Satisfaction Among Health Workers in Deprived Districts of Ghana: A Cross-Sectional Study

**DOI:** 10.1101/2025.09.04.25335137

**Authors:** Kojo Mensah Sedzro, Leonard Baatiema, Kassim Basit, William Akatoti, Sam Amon, India Hotopf, Joanna Raven, Patricia Akweongo

## Abstract

**Background:** Job satisfaction among health workers is critical for improving healthcare delivery and staff retention. This study investigated employment-related determinants of job satisfaction among health workers in three deprived districts of Ghana.

**Methods:** We conducted a cross-sectional survey of 200 public-sector health workers in Kwahu Afram Plains North, Kwahu Afram Plains South, and Kwahu East. A structured questionnaire captured socio-demographic characteristics, employment variables, and satisfaction across domains including teamwork, rewards, communication, and working conditions. Descriptive statistics and non-parametric tests assessed group differences. Ordered logistic regression identified predictors of two outcomes: willingness to recommend the facility as a workplace and overall career satisfaction at the facility.

**Results:** Participants were predominantly female (67.5%) and on permanent contracts (98.7%); median age was 32 years. District, age group, and contract type showed significant differences in satisfaction. Compared with Afram Plains North, working in Kwahu East was associated with higher career satisfaction (AOR=2.70; 95% CI: 1.14–6.41). Older age strongly predicted career satisfaction: workers ≥40 years had higher odds than those aged 20–29 (AOR=13.56; 95% CI: 3.71–49.56). Permanent employment was associated with lower satisfaction relative to temporary contracts (career satisfaction AOR=0.05; 95% CI: 0.01–0.44). Satisfaction with rewards (AOR=4.35; 95% CI: 2.23– 8.50) and communication (facility recommendation AOR=1.98; 95% CI: 1.23–3.19) were pivotal positive determinants. Sex and marital status were not significantly associated with satisfaction.

**Conclusions:** In Ghana’s deprived districts, comprehensive reward systems and effective workplace communication are central to improving job satisfaction and retention. The inverse association between permanent contracts and satisfaction suggests a need to review employment terms and career development pathways for rural posts. Tailored, context-specific interventions addressing rewards, communication, and progression opportunities may enhance morale and support sustained service delivery in underserved areas.

## Introduction

Global shortages and maldistribution of health workers have been identified as major obstacles to achieving universal health coverage by 2030 [1] Recent estimates indicate a worldwide shortfall of about 15 million health workers in 2020 and this is projected to decrease to about 10 million by 2030 [1]. Despite these shortfalls, low-middle income countries suffer the brunt of these workforce shortages especially in rural and deprived areas [3,4].

Ensuring a well-motivated and equitably distributed health workforce has therefore become a global priority. For instance, the World Health Organization recommends targeted strategies to develop, recruit and retain healthcare providers in rural and hard to reach areas, including training more practitioners from rural backgrounds, improving rural training exposure and establishing a clear career development pathway for those serving in deprived areas [4].

In line with these goals, the Ghana’s national health policies emphasize on the equitable deployment and retention of qualified health professionals across all regions. The Ministry of Health envisions a health workforce that is adequately staffed, highly motivated and committed to delivering quality care nationwide [4].

Thus, recent initiatives such as the Ghana Public Health Workforce Strategic Plan (2022-207) seeks address persistent challenges in health worker recruitment, distribution, development, motivation and retention in order to meet universal health coverage by 2030 [5]. Thus, addressing issues of job satisfaction among health workers has been earmarked as fundamental [6–8]. Job satisfaction among health workers is a cornerstone of effective healthcare delivery, particularly in resource-deprived settings where systemic challenges exacerbate workforce attrition and burnout [9,10]. Globally, studies underscore the critical role of job satisfaction in enhancing service quality, staff retention, and patient outcomes [11]. Further to this, it has been established that communication, rewards, and working conditions are also critical drivers of satisfaction [12,13]. Research in high-income contexts emphasizes the interplay between workplace culture, leadership, and job satisfaction [14,15].

In sub-Saharan Africa, including Ghana, health workers in rural and deprived districts face compounded challenges such as inadequate remuneration, limited career progression, and insufficient resources, all of which undermine morale and performance [16–20]. Existing studies in Ghana highlight the influence of intrinsic and extrinsic motivators, such as recognition and financial incentives, on health worker satisfaction [9,10]. However, these findings often focus on urban settings, overlooking the unique socio-economic and operational realities of rural and deprived districts. For instance, permanent employment contracts which traditionally linked to job security and higher satisfaction in global literature [21,22] may paradoxically foster dissatisfaction in resource-constrained environments due to stagnant career pathways or unmet expectations. Similarly, demographic factors like age and professional qualifications, which shape satisfaction in other contexts [23,24], remain underexplored in Ghana’s rural health sector. Furthermore, while non-monetary incentives like mentorship and professional development are gaining traction globally [25,26], their applicability and impact in deprived districts remain unclear. This study aimed to investigate the socio-demographic and employment-related determinants of job satisfaction among health workers in three deprived districts of Ghana.

The findings of the study aim to inform targeted policies to enhance retention and service delivery in underserved regions, contributing to broader discourse on health workforce motivation in low-resource settings. Insights from this study are anticipated to support Ghana’s efforts to addressing health workforce retention issues towards the attainment of UHC. These lessons may be useful in shaping health workforce policies to address retention in similar contexts in low-middle income countries.

## Materials and methods

### Study Design and Setting

This cross-sectional analytical study was conducted among health workers in three under-resourced districts of Ghana: Kwahu Afram Plains North, Kwahu Afram Plains South, and Kwahu East. These districts were purposively selected due to their socioeconomic challenges, and health workforce challenges which may impact workforce motivation and retention.

### Study Population

The study population consisted of health workers from various professional categories, including physicians, nurses, midwives, pharmacists, allied health workers, and dispensary assistants. All participants were working in public health facilities across the three districts at the time of data collection. The inclusion criteria were: (1) Actively working as a health worker in one of the selected districts and (2) Health workers with a minimum duration of at least one year of employment to ensure they have sufficient experience to provide meaningful insights.

### Sampling and Sample Size

A multistage sampling approach stratified facilities into primary (community clinics) and secondary (district hospitals) care levels. Random selection ensured proportional representation across strata. Using the Cochran formula (confidence level = 95%, margin of error = 5%, anticipated prevalence = 50%), we calculated a sample size of 200 health workers.

### Data Collection

Data were collected using a structured questionnaire. The questionnaire was developed based on previously validated survey instruments used to assess health worker motivation and satisfaction in LMIC settings (Bennet et al 2000, Franco et al 2004, Blaauw et al. 2013, Ahmad et al 2020, Vallieres et al 2021) and was pre-tested for validity and reliability in Ghana before full data collection. The questionnaire was designed to capture information on socio-demographic characteristics (age, sex, marital status, and highest qualification), employment-related variables (professional title, type of contract), and various aspects of job satisfaction (teamwork, supervision, rewards, empowerment and participation, training and individual development, communication, working conditions, and management). The questionnaire included Likert-scale items to measure satisfaction levels across these thematic areas, with responses ranging from “Strongly Disagree” to “Strongly Agree.” The thematic areas assessed in the study were Teamwork, Supervision, Rewards, Empowerment and Participation, Training and Individual Development, Communication, Working Conditions, Management and Overall Satisfaction. These areas were selected based on a review of the literature on job satisfaction among health workers, to provide a comprehensive assessment of factors influencing job satisfaction in the selected districts.

### Data Analysis

Data were analysed using Stata version 18.5. Descriptive statistics were calculated to summarize the socio-demographic and employment-related characteristics of the study participants. The association between these variables and overall job satisfaction was assessed using non-parametric tests due to the ordinal nature of the satisfaction measures. Specifically, the Kruskal-Wallis’s test was used to compare median satisfaction scores across groups defined by categorical variables (e.g., district of employment, sex, age group, marital status, highest qualification, professional title, and employment contract type) and effect size imputed to determine the variation of effect among variables that are significant. Effect sizes were interpreted as follows: η² = 0.01–0.06 (small), 0.06–0.14 (moderate), and >0.14 (large). To further explore the factors associated with job satisfaction, ordered logistic regression models with robust standard errors were employed. These models were used to estimate adjusted odds ratios (AOR) and 95% confidence intervals (CI).

The proportional odds (parallel regression) assumption was evaluated using the likelihood ratio test (LRT) via the omodel command. This test compares the constrained ordered logistic regression model to an unrestricted generalized ordered logit model. For Model 1 (Facility Recommendation), the LRT yielded χ²(39) = 40.97 (*p* = 0.384), and for Model 2 (Career Satisfaction), χ²(39) = 52.02 (*p* = 0.079). Both models satisfied the proportional odds assumption (*p* > 0.05), indicating that the effects of predictors were consistent across all ordinal thresholds of the dependent variables. Consequently, the ordered logistic regression framework was retained for analysis. Variables included in the regression models were selected based on theoretical relevance and the results of bivariate analyses.

### Ethical Considerations

Written informed consent was obtained from all participants prior to their involvement in the study. Participation was voluntary, and participants were assured of the confidentiality and anonymity of their responses. Data was stored securely and accessed only by the research team. Ethical approval was granted by the Ghana Health Service Ethics Review Committee (GHS-ERC: 007/02/24) and Liverpool School of Tropical Medicine (24-002) in the United Kingdom.

## Results

### Demographic Characteristic of study participants

A total of 200 health workers participated in the study, comprising 67 respondents from Kwahu Afram Plains North, 65 from Kwahu Afram Plains South, and 68 from Kwahu East. The demographic profile of the sample is summarized in Table 1. Overall, about two-thirds of the respondents were female 135 (67.5%). The median age of participants was 32 years (IQR: 29-35), with the largest age being 30–39 years (67.5%). Almost all participants were employed on permanent government contracts (98.5%). This reflects the employment structure in the Ghanaian public health sector, where most workers are civil service employees. The distribution of health professional cadres included nurses and midwives as majority. There were some notable differences in participant characteristics across the three districts in sex (*p* = 0.033), age (*p* = 0.015), and highest qualification (*p* = 0.022) (Table 1.0), with Kwahu East having a higher proportion of female health workers and higher proportion of staff with diplomas and fewer with certificates compared to Afram Plains North and South Districts. These baseline differences contextualize the subsequent analysis of job satisfaction.

**Table 1.** Background Characteristics of Participants by districts.

| Demographics | Kwahu<br>Afram<br>Plains<br>North<br>N=67 | Kwahu<br>Afram Plains<br>South<br>N=65 | Kwahu<br>East<br>N=68 | Total<br>N=200 | p-value |
| --- | --- | --- | --- | --- | --- |
| **Sex** |  |  |  |  | **0.033** |
| Male | 27 (40.3) | 24 (36.9) | 14 (20.6) | 65 (32.5) |  |
| Female | 40 (59.7) | 41 (63.1) | 54 (79.4) | 135 (67.5) |  |
| **Age (In Years)** |  |  |  |  |  |
| **Median (IQR)** | 32 (30-35) | 30 (29-33) | 32 (31-36) | 32 (29-35) | **0.015** |
| 20-29 | 16 (23.9) | 23 (35.4) | 12 (17.6) | 51 (25.5) |  |
| 30-39 | 43 (64.2) | 41 (63.1) | 51 (75.0) | 135 (67.5) |  |
| 40+ | 8 (11.9) | 1 (1.5) | 5 (7.4) | 14 (7.0) |  |
| **Marital Status** |  |  |  |  | 0.052 |
| Currently married or living together | 26 (38.8) | 28 (43.1) | 41 (60.3) | 95 (47.5) |  |
| Divorced/Separated/ Widowed | 0 (0.0) | 1 (1.5) | 4(4.4) | 4 (2.0) |  |
| Never married | 41 (61.2) | 36 (55.4) | 24 (35.3) | 101 (50.5) |  |
| **Highest qualification** |  |  |  |  | **0.022** |
| Certificate | 33 (49.3) | 36 (55.4) | 27 (39.7) | 96 (48.0) |  |
| Diploma | 16 (23.9) | 21 (32.3) | 33 (48.5) | 70 (35.0) |  |
| 1st degree (Bachelor) | 13 (19.4) | 7 (10.8) | 8 (11.8) | 28 (14.0) |  |
| 2nd degree (Master's) | 2 (3.0) | 1 (1.5) | 0 (0.0) | 3 (1.5) |  |
| Other | 3 (4.5) | 0 (0.0) | 0 (0.0) | 3 (1.5) |  |
| **Year of completion** |  |  |  |  | **0.086** |
| 2004-2014 | 5 (7.5) | 1 (1.5) | 10 (14.7) | 16 (8.0) |  |
| 2015-2019 | 46 (68.7) | 50 (76.9) | 44 (64.7) | 140 (70.0) |  |
| 2020-2024 | 16 (23.9) | 14 (21.5) | 14 (20.6) | 44 (22.0) |  |
| **Professional Title** |  |  |  |  | **0.683** |
| Physician/Assistant Medical Officer | 2 (3.0) | 3 (4.6) | 2 (2.9) | 7 (3.5) |  |
| Nurse/Nursing Assistant | 27 (40.3) | 30 (46.2) | 22 (32.4) | 79 (39.5) |  |
| Community Nurse/Mental Health Officer | 17 (25.4) | 13 (20.0) | 21 (30.9) | 51 (25.5) |  |
| Midwife | 11 (16.4) | 13 (20.0) | 14 (20.6) | 38 (19.0) |  |
| Pharmacist/Dispensary Assistant | 6 (9.0) | 5 (7.7) | 8 (11.8) | 19 (9.5) |  |
| Allied Health Worker | 4 (6.0) | 1 (1.5) | 1 (1.5) | 6 (3.0) |  |
| **Permanent contract of employment** | 65 (97.0) | 65 (100.0) | 67 (98.5) | 197 (98.5) | **0.658** |

Data are presented as median (IQR) for continuous measures, and n (%) for categorical measures.

### Associations Between Variables and Job Satisfaction Domains

The bivariate analyses examined how various socio-economic and job-related factors were associated with health workers’ satisfaction across the key thematic domains. Table 2 and 3 summarizes these associations. The district where health workers served showed significant association with certain satisfaction domains. District of employment was significantly associated with teamwork (p = 0.016, η² = 0.04), working conditions (p < 0.001, η² = 0.11), and overall satisfaction (p < 0.001, η² = 0.10). Age group showed a significant relationship with empowerment (p = 0.004, η² = 0.05), with older health workers reporting higher satisfaction levels. Health workers in Kwahu East tended to report slightly higher teamwork satisfaction scores compared to those in the Afram Plains districts, although the effect size for teamwork was small. For working conditions, the difference was more pronounced (moderate size effect) where respondents in Kwahu East indicated better satisfaction with their workload, infrastructure and resources than the other two districts, which aligns with anecdotal reports that Kwahu East is relatively better resourced.

**Table 2.** Associations between socio-demographic and employment-related variables and satisfaction domains (teamwork, supervision, reward, empowerment and participation).

| Demographics | Teamwork |  | Supervision |  | Reward |  | Empowerment and Participation |  |
| --- | --- | --- | --- | --- | --- | --- | --- | --- |
| | Median [IQR] | P-value/ $\eta^2$ | Median [IQR] | P-value/ $\eta^2$ | Median [IQR] | P-value/ $\eta^2$ | Median [IQR] | P-value/ $\eta^2$ |
| **District** |  | 0.016/0.04 |  | 0.364 |  | 0.052 |  | 0.521 |
| Kwahu Afram Plains North | 4.40[4.00,4.80] |  | 4.00[3.71,4.43] |  | 2.86[2.71,3.29] |  | 4.00[3.80,4.40] |  |
| Kwahu Afram Plains South | 4.20[4.00,4.60] |  | 4.00[3.57,4.29] |  | 2.71[2.43,3.14] |  | 4.00[3.80,4.40] |  |
| Kwahu East | 4.10[4.00,4.40] |  | 4.00[3.57,4.07] |  | 2.86[2.43,3.29] |  | 4.00[3.80,4.40] |  |
| **Sex** |  | 0.53 |  | 0.159 |  | 0.061 |  | 0.03/0.02 |
| Male | 4.20[4.00,4.80] |  | 3.86[3.43,4.14] |  | 3.00[2.57,3.29] |  | 4.20[3.80,4.60] |  |
| Female | 4.20[4.00,4.60] |  | 4.00[3.71,4.14] |  | 2.71[2.43,3.14] |  | 4.00[3.60,4.40] |  |
| **Age-group** |  | 0.626 |  | 0.343 |  | 0.66 |  | 0.004/0.05 |
| 20-29 | 4.20[4.00,4.60] |  | 3.86[3.43,4.14] |  | 2.71[2.43,3.29] |  | 3.80[3.60,4.20] |  |
| 30-39 | 4.40[4.00,4.75] |  | 4.00[3.71,4.14] |  | 2.86[2.57,3.29] |  | 4.00[3.80,4.40] |  |
| 40+ | 4.40[4.00,4.60] |  | 4.07[4.00,4.29] |  | 2.79[2.43,3.00] |  | 4.40[4.00,4.60] |  |
| **Marital Status** |  | 0.305 |  | 0.945 |  | 0.817 |  | 0.872 |
| Currently married or living together | 4.20[4.00,4.60] |  | 4.00[3.71,4.14] |  | 2.86[2.57,3.14] |  | 4.00[3.80,4.40] |  |
| Divorced/Separated | 4.80[4.00,5.00] |  | 3.86[3.43,4.14] |  | 2.57[2.43,3.14] |  | 4.40[3.80,4.60] |  |
| Widowed | 4.40[4.40,4.40] |  | 4.14[4.14,4.14] |  | 2.57[2.57,2.57] |  | 4.00[4.00,4.00] |  |
| Never married | 4.40[4.00,4.60] |  | 3.86[3.57,4.14] |  | 2.86[2.43,3.29] |  | 4.00[3.80,4.40] |  |
| **Highest qualification** |  | 0.738 |  | 0.344 |  | <0.001/0.09 |  | 0.651 |
| Certificate | 4.20[4.00,4.70] |  | 4.00[3.71,4.14] |  | 3.00[2.57,3.29] |  | 4.00[3.60,4.40] |  |
| Diploma | 4.20[4.00,4.60] |  | 4.00[3.57,4.14] |  | 2.57[2.14,2.86] |  | 4.00[3.80,4.20] |  |
| 1st degree (Bachelor) | 4.40[4.00,4.60] |  | 4.00[3.64,4.5] |  | 2.86[2.64,3.14] |  | 4.30[3.8,4.60] |  |
| 2nd degree (Master's) | 4.00[2.40,4.40] |  | 1.43[1.00,4.00] |  | 3.14[2.14,4.14] |  | 4.00[4.00,4.40] |  |
| Other | 4.00[4.00,4.60] |  | 4.00[3.86,4.57] |  | 3.14[2.71,3.57] |  | 4.00[3.80,4.80] |  |
| **Professional Title** |  | 0.134 |  | 0.142 |  | 0.214 |  | 0.791 |
| Physician/Assistant Medical Officer | 4.20[3.20,4.40] |  | 3.14[1.29,4.00] |  | 2.86[2.29,3.14] |  | 4.00[4.00,4.40] |  |
| Nurse/Nursing Assistant | 4.20[4.00,4.80] |  | 4.00[3.57,4.29] |  | 2.86[2.43,3.29] |  | 4.00[3.80,4.40] |  |
| Community Nurse/Mental Health Officer | 4.20[4.00,4.40] |  | 3.86[3.71,4.14] |  | 2.86[2.57,3.29] |  | 4.00[3.60,4.40] |  |
| Midwife | 4.40[4.00,4.80] |  | 4.00[3.86,4.29] |  | 2.64[2.14,3.00] |  | 4.00[3.80,4.40] |  |
| Pharmacist/Dispensary Assistant | 4.00[4.00,4.40] |  | 4.00[3.43,4.00] |  | 3.14[2.57,3.29] |  | 4.00[3.80,4.40] |  |
| Allied Health Worker | 4.4[3.40,4.60] |  | 4.36[3.71,4.57] |  | 2.86[2.57,2.86] |  | 4.20[3.60,4.80] |  |
| Permanent contract of employment 1 | 4.20[4.00,4.60] | 0.461 | 4.00[3.57,4.14] | 0.859 | 2.86[2.43,3.29] | 0.801 | 4.00[3.80,4.40] | 0.742 |

Values are median [IQR]. P-values from Kruskal–Wallis tests. $\eta^2$ = effect size (0.01–0.06 = small; 0.06–0.14 = moderate; >0.14 = large).

**Table 3.** Associations between socio-demographic and employment-related variables and satisfaction domains (training and individual development, communication, working conditions, management) and overall satisfaction.

| Demographics | Training and Individual Development |  | Communication |  | Working Conditions |  | Management |  | Overall |  |
| --- | --- | --- | --- | --- | --- | --- | --- | --- | --- | --- |
| | Median [IQR] | P-value/$\eta^2$ | Median [IQR] | P-value/$\eta^2$ | Median [IQR] | P-value/$\eta^2$ | Median [IQR] | P-value/$\eta^2$ | Median [IQR] | P-value/$\eta^2$ |
| **District** |  | 0.242 |  | 0.719 |  | <0.001/0.11 |  | 0.948 |  | <0.001/0.10 |
| Kwahu Afram Plains North | 4.00[3.43,4.29] |  | 3.80[3.40,4.00] |  | 3.17[2.83,3.50] |  | 3.50[3.00,4.00] |  | 2.50[1.50,3.00] |  |
| Kwahu Afram Plains South | 4.00[3.57,4.14] |  | 3.60[3.40,4.00] |  | 3.00[2.50,3.17] |  | 3.50[3.00,4.00] |  | 1.50[1.00,2.50] |  |
| Kwahu East | 3.79[3.36,4.07] |  | 3.60[3.10,4.00] |  | 3.42[3.00,3.67] |  | 3.50[3.00,4.00] |  | 2.50[2.00,3.25] |  |
| **Sex** |  | 0.068 |  | 0.902 |  | 0.934 |  | 0.756 |  | 0.162 |
| Male | 4.00[3.57,4.14] |  | 3.60[3.40,4.20] |  | 3.25[2.58,3.58] |  | 3.50[3.00,4.00] |  | 2.00[1.50,2.50] |  |
| Female | 3.71[3.29,4.00] |  | 3.60[3.20,4.00] |  | 3.17[2.83,3.50] |  | 3.50[3.00,4.00] |  | 2.50[1.50,3.00] |  |
| **Age-group** |  | 0.071 |  | 0.758 |  | 0.156 |  | 0.374 |  | 0.067 |
| 20-29 | 3.71[3.29,4.14] |  | 3.60[3.20,4.00] |  | 3.17[2.83,3.67] |  | 4.00[3.00,4.00] |  | 2.50[1.50,3.00] |  |
| 30-39 | 3.86[3.43,4.00] |  | 3.60[3.20,4.00] |  | 3.17[2.67,3.50] |  | 3.50[3.00,4.00] |  | 2.50[1.00,3.00] |  |
| 40+ | 4.00[4.00,4.43] |  | 4.00[3.60,4.00] |  | 3.42[2.92,4.08] |  | 3.75[3.00,4.00] |  | 3.00[1.50,4.50] |  |
| **Marital Status** |  | 0.233 |  | 0.598 |  | 0.805 |  | 0.099 |  | 0.308 |
| Currently married or living together | 3.86[3.29,4.14] |  | 3.60[3.20,4.00] |  | 3.17[2.83,3.50] |  | 3.00[3.00,4.00] |  | 2.50[1.50,3.00] |  |
| Divorced/Separated | 3.71[3.57,4.00] |  | 3.8[3.80,4.20] |  | 2.58[2.08,4.33] |  | 3.00[3.00,3.50] |  | 3.00[1.00,4.50] |  |
| Widowed | 3.14[3.14,3.14] |  | 4.00[4.00,4.00] |  | 2.92[2.92,2.92] |  | 3.00[3.00,3.00] |  | 5.00[5.00,5.00] |  |
| Never married | 3.86[3.57,4.14] |  | 3.60[3.20,4.00] |  | 3.17[2.75,3.58] |  | 4.00[3.00,4.00] |  | 2.50[1.50,3.00] |  |
| **Highest qualification** |  | 0.017/0.06 |  | 0.39 |  | 0.416 |  | 0.682 |  | 0.266 |
| Certificate | 3.93[3.57,4.14] |  | 3.8[3.40,4.20] |  | 3.12[2.83,3.54] |  | 3.50[3.00,4.00] |  | 2.50[1.00,3.00] |  |
| Diploma | 3.79[3.29,4.00] |  | 3.60[3.20,4.00] |  | 3.21[2.67,3.58] |  | 3.75[3.00,4.00] |  | 2.50[1.50,3.00] |  |
| 1st degree (Bachelor) | 3.64[3.43,4.00] |  | 3.60[2.90,4.00] |  | 3.21[2.83,3.42] |  | 3.00[3.00,4.00] |  | 2.00[1.00,2.50] |  |
| 2nd degree (Master's) | 4.29[3.43,4.57] |  | 3.60[1.80,4.20] |  | 2.58[2.00,4.08] |  | 3.00[1.50,4.50] |  | 2.00[1.00,3.50] |  |
| Other | 4.14[3.57,4.43] |  | 3.60[3.60,4.00] |  | 3.67[3.50,3.83] |  | 4.00[3.00,4.00] |  | 3.00[2.50,4.00] |  |
| **Professional Title** |  | 0.428 |  | 0.682 |  | 0.043/0.06 |  | 0.241 |  | 0.139 |
| Physician/Assistant Medical Officer | 3.43[3.43,3.71] |  | 3.60[3.00,4.00] |  | 2.58[2.50,2.92] |  | 3.00[3.00,4.00] |  | 1.50[1.00,2.00] |  |
| Nurse/Nursing Assistant | 3.86[3.29,4.14] |  | 3.60[3.20,4.00] |  | 3.17[2.83,3.42] |  | 3.00[3.00,4.00] |  | 2.50[1.00,3.00] |  |
| Community Nurse/Mental Health Officer | 3.86[3.29,4.14] |  | 3.8[3.40,4.20] |  | 3.17[2.75,3.58] |  | 3.50[3.00,4.00] |  | 2.50[1.50,3.00] |  |
| Midwife | 3.86[3.43,4.00] |  | 3.8[3.20,4.00] |  | 3.17[2.67,3.58] |  | 4.00[3.00,4.00] |  | 2.50[1.50,3.00] |  |
| Pharmacist/Dispensary Assistant | 4.00[3.71,4.14] |  | 3.4[2.60,4.00] |  | 3.50[2.42,3.67] |  | 3.50[3.00,4.00] |  | 2.50[1.50,3.50] |  |
| Allied Health Worker | 4.14[4.00,4.43] |  | 3.60[3.40,3.80] |  | 3.58[3.17,4.25] |  | 3.75[3.00,4.50] |  | 1.50[1.50,1.50] |  |
| Permanent contract of employment <sup>1</sup> | 3.86[3.43,4.14] | 0.28 | 3.60[3.20,4.00] | 0.513 | 3.17[2.75,3.50] | 0.9 | 3.50[3.00,4.00] | 0.055 | 2.50[1.50,3.00] | 0.044 |

Values are median [IQR]. P-values from Kruskal–Wallis tests. $\eta^2$ = effect size (0.01–0.06 = small; 0.06–0.14 = moderate; >0.14 = large)..

Significant associations were also observed between highest qualification and satisfaction with rewards (p < 0.001, η² = 0.09) and training and individual development (p = 0.017, η² = 0.06). Professional title was significantly associated with satisfaction related to working conditions (p = 0.043, η² = 0.06). Additionally, contract type was significantly associated with overall satisfaction (p = 0.044, η² = 0.03), where temporary staff reported higher median scores.

Most effect sizes were small (η² < 0.06), with moderate effect sizes observed for working conditions and rewards, indicating practical relevance of these domains.

### Predictive Factors for Facility Recommendation and Career Satisfaction among Health Workers

To identify independent determinants of job satisfaction, two ordered logistic regression models estimated. Model 1 estimated the likelihood of recommending the health facility to others to work in as measure of overall satisfaction and model 2 estimated career satisfaction where the participants are happy to spend their careers at that health facility (Table 4).

**Table 4:**
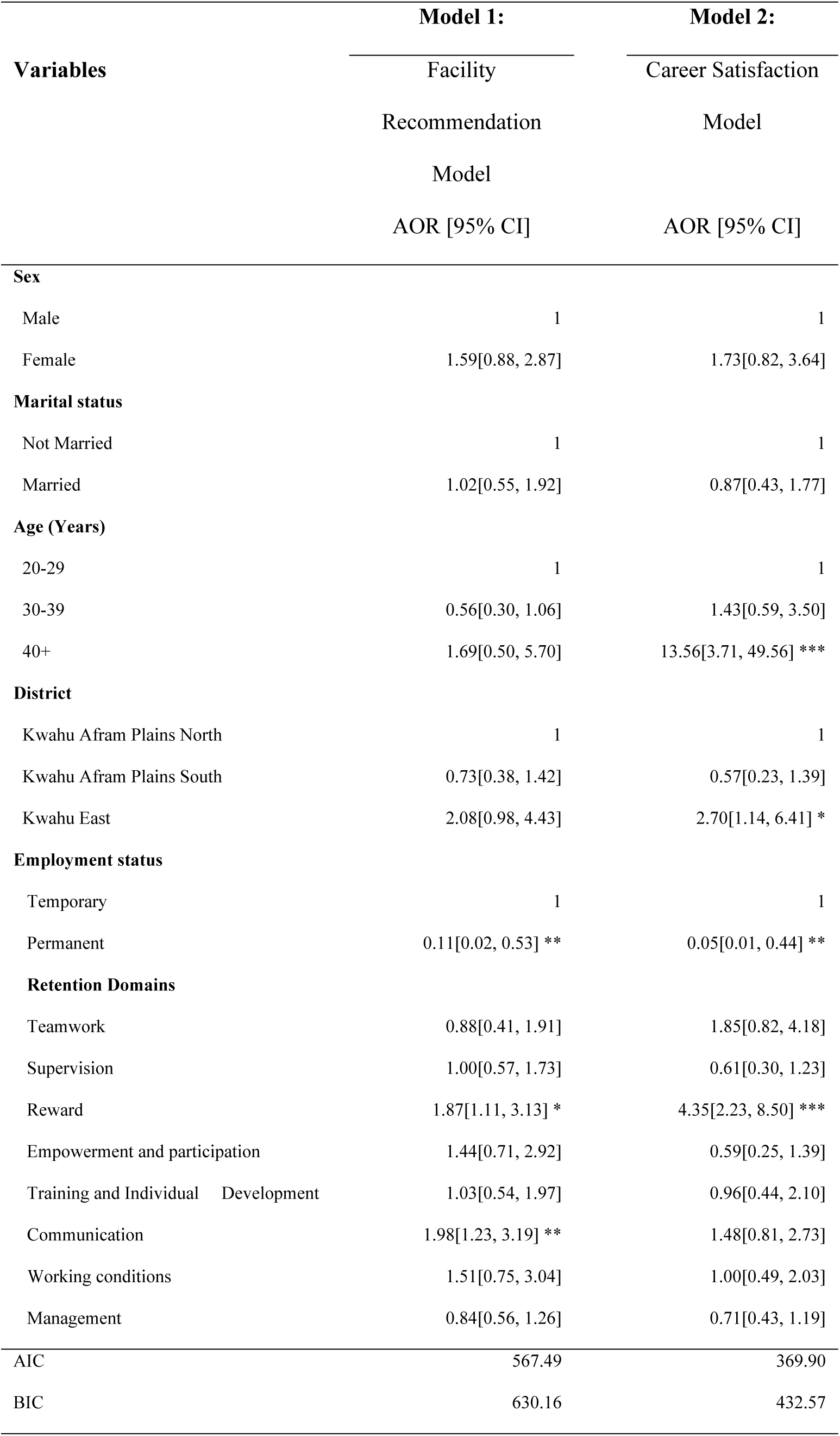
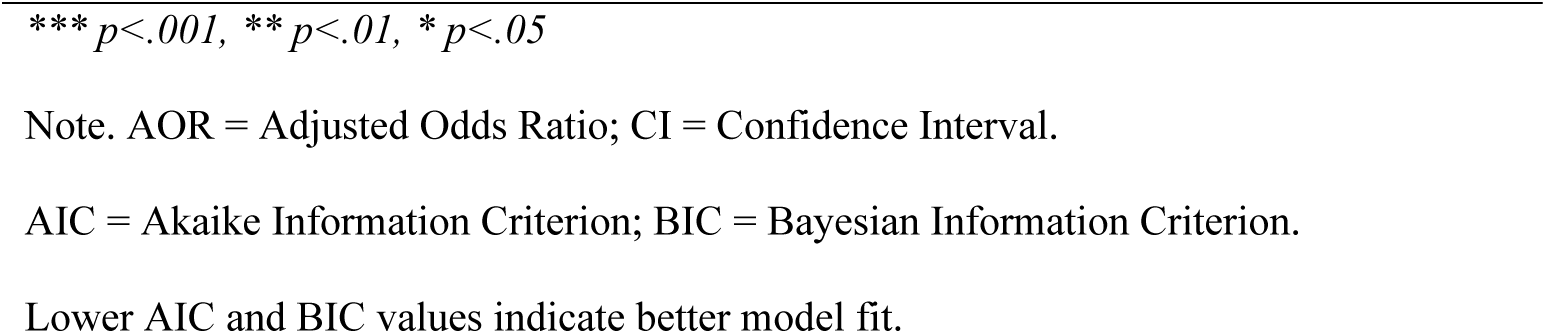
Factors Affecting Health Workers’ Facility Recommendation and Career Satisfaction.

Key statistically significant predictors of facility recommendation included satisfaction with rewards (AOR = 1.87; 95% CI: 1.11–3.13; p < 0.05) and communication (AOR = 1.98; 95% CI: 1.23–3.19; p < 0.01). Satisfaction with rewards was a strong predictor in both models (facility recommendation) with higher satisfaction with the rewards system associated with greater odds of recommending the facility. The effect of career satisfaction was even larger indicating that those who felt adequately rewarded were four times more likely to express satisfaction with a long-term career at the facility. Having a permanent contract, was however associated with lower odds (AOR = 0.11; 95% CI: 0.02–0.53; p < 0.01). For career satisfaction, significant predictors were older age (≥40 years) (AOR = 13.56; 95% CI: 3.71–49.56; p < 0.001), satisfaction with rewards (AOR = 4.35; 95% CI: 2.23–8.50; p < 0.001). After adjusting for district employment (Kwahu East) (AOR = 2.70; 95% CI: 1.14–6.41; p < 0.05), working in Kwahu East remained associated with higher job satisfaction compared to Afram Plains North. Similarly, staff in Kwahu East had about 2.7 times odds of being satisfied with their career at their facility relative to those in the Afram Plains North. Working in Afram Plains South was not significantly different from Afram Plains North suggesting that Kwahu East had better resources and community conditions that foster higher satisfaction beyond the individual level factors. Having a permanent contract remained a significant determinant (AOR = 0.05; 95% CI: 0.01–0.44; p < 0.01).

## Discussion

This study examined the drivers of job satisfaction among health workers in three deprived districts of Ghana and provides insights into how both personal and job-related factors influence health worker satisfaction in resource-constrained settings. The findings highlight several critical determinants, such as roles of employment conditions, workplace environment and the implications for health work policies in Ghana and similar low resource contexts.

One key finding of this study is the negative association between having a permanent job contract and job satisfaction. Health workers on permanent employment contracts had significantly lower odds of recommending their facility or expressing long-term career satisfaction, even though permanent employment is typically associated with greater job security. This result suggests that in these deprived districts, permanent staff may experience unmet expectations or frustrations related to limited opportunities for promotion, slow salary progression, or bureaucratic constraints which tend to outweigh the benefits of job security. It is possible that once workers obtain a permanent employment contract, they become aware of the challenges in these deprived areas such as lack of resources or stagnation in rural postings and hence feel “stuck,” whereas those on temporary contracts might still anticipate future mobility or improvement. While we observed lower satisfaction among permanent workers, the near-total absence of temporary staff in our sample (98.5% permanent) necessitates caution in interpreting this finding. Future studies with balanced representation should verify whether contract type itself drives dissatisfaction or whether it proxies for other factors (e.g., tenure length or unmet expectations).

Similar observations have been reported in other contexts. For example, a study in rural Ghana noted that health workers often value career development prospects as much as job security, and when permanency comes without growth opportunities, it can diminish motivation [27]. Contrary to studies linking permanent contracts to job security [28] our findings suggest dissatisfaction may stem from inflexible terms or stagnation in career growth. This finding suggests that permanent contracts may come with unmet expectations, job dissatisfaction, or a perceived lack of career advancement opportunities. On the contrary, some studies have reported that workers with permanent contracts tend to account for higher job satisfaction due to increased job security and benefits, which can enhance motivation and reduce turnover intentions [21,28,29]. This underscores the need for health sector policymakers to re-examine the terms and conditions of permanent employment posts in underserved areas. Ensuring that rural permanent staff have clear career pathways, regular promotions, and continuing education opportunities could mitigate the dissatisfaction observed.

Studies show that a positive workplace culture characterised by inclusivity boosts employee engagement and job satisfaction, which correlates with a higher likelihood of recommending the workplace of health workers [14,25]. Leadership also plays a crucial role, as effective leadership fosters a supportive environment and encourages health workers to speak positively about their organisation, a behaviour often referred to as employee advocacy [14]. This advocacy reflects a worker’s willingness to recommend their employer, defend the organisation’s reputation, or attract potential recruits. However, high levels of perceived stress negatively impact health workers’ willingness to recommend their employer or place of work. Factors such as value correspondence, leadership communication, and work schedule flexibility can mitigate this stress [11]. Moreover, strong leadership and a positive workplace culture can foster better staff–patient relationships and quality of care, leading to more rewarding interactions. These positive interactions with patients and satisfaction with care delivery, in turn, have been shown to enhance health workers’ likelihood of recommending their workplace[13].

In this study, age and experience emerged as significant predictors of job satisfaction, with health workers aged ≥40 reporting substantially higher satisfaction than younger colleagues. This aligns with prior evidence suggesting that longer job tenure enables older workers to develop stronger workplace attachment through accumulated experience, greater role stability, and alignment of expectations with professional realities [24,30] As workers gain seniority, they typically attain positions of greater responsibility, refine coping mechanisms, and establish community or family ties that reinforce contentment and retention.

By comparison, the youngest cohort (20–29 years) expressed the lowest satisfaction level, a finding potentially explained by unmet career expectations, developmental needs, or dissatisfaction with entry-level conditions [23]. Unlike studies noting patterns where both youngest and oldest workers report high satisfaction [31] our context reveals a linear progression where satisfaction increases with age. This suggests that younger health workers’ aspirations for advancement or alternative opportunities may amplify their readiness to seek new roles, while their older counterparts’ investment in stability and established roles fosters deeper commitment.

The lack of significant difference between men and women in overall satisfaction suggests that the harsh working conditions in deprived districts affect all staff similarly. While some research evidence from higher-income contexts indicates women sometimes report higher job satisfaction in healthcare roles ([23],) in our study both genders faced the same systemic challenges (resource shortages, heavy workloads). This suggests that systemic challenge such as resource constraints and workload may uniformly affect job satisfaction across genders in deprived districts, diminishing sex-based differences observed in other contexts [32]. This finding highlights that efforts to improve job satisfaction in such settings should benefit all workers and that gender-specific interventions might not be a primary concern compared to broader improvements in the work environment.

The workplace environment factors stood out as critical. Satisfaction with rewards which encompassing salary, allowances, and recognition was the strongest predictor of both outcomes. This highlights the importance of effective reward systems, including financial incentives, recognition, and career development opportunities, in enhancing job satisfaction and retention. Studies show that extrinsic motivation (e.g., financial rewards) significantly predicts job satisfaction, but this relationship is often strengthened when intrinsic motivation (such as a sense of personal fulfilment or professional growth) is also present. In this context, intrinsic motivation may act as a mediator, helping to explain how and why external incentives lead to improved satisfaction [12]. Motivation strategies that focus on non-financial incentives like motivating personnel for excellence and prioritizing the needs of staff can also yield high satisfaction levels among health staff [26].

This reinforces longstanding evidence that adequate compensation is fundamental to motivating health workers to remain in underserved areas [33]. Ghana, like many LMICs, has struggled with health worker retention in rural posts due in part to perceived financial disadvantages of working in remote areas. The strong association we found suggests that improving financial and non-financial reward systems could substantially boost morale. Non-financial rewards such as recognition programs, employee-of-the-month awards, opportunities for professional development, and clear promotion criteria can complement salary increases. These can fulfill health workers’ intrinsic needs for appreciation and growth, which, alongside extrinsic rewards, contribute to overall job satisfaction. Our results reflect findings from other studies in Ghana that echo that both extrinsic incentives (like hardship allowances or housing for rural staff) and intrinsic motivators (like a sense of accomplishment and community impact) are necessary to keep health workers satisfied and committed to their posts [33]. For example, a meta-analysis of factors associated with health worker satisfaction found that remuneration, recognition, and career advancement were consistently linked to higher satisfaction across diverse settings (Rizky et al., 2023). In Ghana’s context, the government has periodically introduced financial incentives for rural service such as additional allowances or salary uplifts for postings in remote areas and our findings provide empirical support that such measures are indeed crucial. However, they should be sustained and coupled with other improvements to have lasting impact.

Effective communication within health facilities fosters transparency, trust, and a positive work environment contributing to higher job satisfaction and willingness to recommend the workplace to others [34]. In our study areas, where resources are scarce, clear communication can also help manage expectations and reduce frustration (for example, explaining reasons for shortages or involving staff in problem-solving). The importance of communication is supported by global evidence that participatory management and open communication channels improve job satisfaction and retention. A study of nurses in Ethiopia, for instance, showed that leadership communication and feedback were associated with better job satisfaction and lower intent to leave [35]. In the Ghana context, deprived districts, fostering a culture of open dialogue between health workers and supervisors or district health managers could alleviate feelings of neglect and improve morale, even when material resources are limited.

It is important to highlight that some factors we hypothesized to influence satisfaction did not show independent effects in the multivariate analysis once rewards and communication were accounted for. Teamwork, supervision quality, working conditions, and management style all had positive correlations with satisfaction in bivariate analysis, but they did not emerge as significant in the final model. This may be due to multicollinearity where facilities with good communication might also have good teamwork and management, making it hard to disentangle their effects with our sample size. Nonetheless, these factors are clearly interrelated parts of the work environment. Strong supportive supervision, for instance, often improves communication and provides recognition (a form of reward), thereby indirectly boosting satisfaction. Similarly, decent working conditions (adequate supplies, manageable workload) can reduce stress and improve staff relationships. This implies that a holistic approach is needed in that while addressing salaries and communication, health system managers should not ignore improvements in supervision, workload management, and facility conditions. Even if these did not show up as independent predictors, they form the context in which rewards and communication are delivered. Other studies in Ghana have found that health workers frequently cite heavy workloads, lack of equipment, and poor living conditions as reasons for dissatisfaction and leaving rural posts [33]. Supervision plays a critical role as health workers receiving adequate supervision are more likely to report job satisfaction motivating health workers to perform better [35,36]. Positive working conditions, including manageable workloads, supportive environments, and empowerment through professional development opportunities also contribute to higher satisfaction levels [37]. These aspects collectively contribute to the work environment and employee morale, and any improvements in these areas can potentially enhance overall job satisfaction. Therefore, tackling those issues remain important for a sustainable retention strategy.

## Limitations

The cross-sectional design limits our ability to draw causal conclusions. It is possible that inherently more motivated or satisfied individuals were more likely to remain in these jobs reflecting the selection effect, rather than the job features alone causing satisfaction or dissatisfaction. Importantly, our sample consisted overwhelmingly of permanent workers (98.5%), which introduces significant selection bias. This imbalance limits the generalizability of findings about contract types and may distort the observed negative association between permanent contracts and satisfaction. We also relied on self-reported measures of satisfaction, which can be influenced by transient moods or social desirability bias. However, the anonymous survey administration was intended to encourage honesty. Another limitation is that our study was confined to three districts within one region of Ghana. Thus, while these are illustrative of deprived rural settings, the results may not generalize to all regions of Ghana or other countries without caution. We did not collect detailed qualitative data that might explain why permanent staff felt dissatisfied, for instance. Future research could complement these findings with qualitative interviews to understand the motivations and frustrations of health workers in such settings.

## Conclusion

This study identified key factors that influence health worker job satisfaction in deprived districts of Ghana. Ensuring fair rewards, effective communication, and career development opportunities for health workers, while addressing the unique challenges tied to employment terms and rural postings are essential steps toward improving retention in deprived areas of Ghana. Also investing in the well-being and satisfaction of its health workforce through both policy reforms and resource allocation, Ghana can make progress towards closing the urban-rural gap in health services. These findings contribute to the wider understanding that a motivated health workforce is the backbone of any health system, especially in low-resource settings. Thus, efforts to continue to address health worker shortages, focusing on job satisfaction and working conditions would improve staff morale and lead to achieving universal health coverage.

### Implications for Policy and Practice

The consistent finding that permanent contracts are associated with lower job satisfaction suggests a need to re-evaluate the terms and conditions of permanent employment. Engaging with permanently contracted health workers to identify their specific concerns and developing targeted interventions can help address these issues and improve satisfaction. The lower odds of career satisfaction among younger health workers highlight the importance of targeted support and development programs for this group. Mentorship programs, career progression opportunities, and initiatives to enhance job satisfaction and retention among younger staff are essential. If younger workers see a pathway to advancement and feel supported, they may be less inclined to leave these districts or the public sector entirely. The significant district differences in job satisfaction suggest that strategies to improve job satisfaction might need to be tailored to each district’s realities. Context-specific interventions can help ensure that interventions are effective and relevant.

## Author Contribution

**Conceptualization:** JR, PA

**Methodology:** JR, PA

**Formal analysis:** KMS

**Data curation:** KMS

**Investigation:** SA, WA

**Project administration:** KB

**Supervision:** KB, WA

**Writing – original draft:** KMS

**Writing – review & editing:** BL, PA, SA, IH

**Funding acquisition:** JR, PA

All authors have read and approved the final manuscript

## Initials map

SKM = Kojo Mensah Sedzro; BL = Baatiema Leonard; KB = Kassim Basit; WA = William Akatoti; SA = Sam Amon; JR = Joanna Raven; PA = Patricia Akweongo.

## Acknowledgement

We are deeply grateful to the health workers who participated in this study and to the facility managers and staff across Kwahu Afram Plains North, Kwahu Afram Plains South, and Kwahu East for their cooperation and support. We thank the District Health Management Teams for facilitation and guidance, and the field supervisors, research assistants, and data entry personnel for their diligent work during data collection and processing. We also appreciate the administrative support provided by colleagues at the School of Public Health, University of Ghana; the Ghana Health Service Eastern region directorate, University of Ghana; and the Liverpool School of Tropical Medicine. Finally, we acknowledge the Ghana Health Service Ethics Review Committee (GHS-ERC: 007/02/24) and the Liverpool School of Tropical Medicine Research Ethics Committee (24-002) for ethical oversight.

## Data Availability

All relevant data are within the paper and its Supporting Information files. The de-identified dataset underlying the findings is provided as S1 Data.

